# Optimal Deployment of Automated External Defibrillators in a Long and Narrow Environment

**DOI:** 10.1101/2022.02.04.22270427

**Authors:** Chih-Hao Lin, Kuan-Chao Chu, Jung-Ting Lee, Chung-Yao Kao

## Abstract

**Aim of the study:** Public access to automated external defibrillators (AEDs) plays a key role in increasing survival outcomes for patients with out-of-hospital cardiac arrest. Based on the concept of maximizing “rescue benefit” of AEDs, we aimed to develop strategies for optimal deployment of AEDs for long and narrow spaces.

**Methods:** We classified the effective coverage of an AED in hot, warm, and cold zones. The AEDs were categorized, according to their accessibility, as fixed, summonable, or patrolling types. The overall rescue benefit of the AEDs were evaluated by the weighted size of their collective hot zones. The optimal strategies for the deployment of AEDs were derived mathematically and numerically verified by computer programs.

**Results:** To maximize the overall rescue benefit of the AEDs, the AEDs should avoid overlapping with each other’s coverage as much as possible. Specific rules for optimally deploying one, two, or multiple AEDs, and various types of AEDs are summarized and presented.

**Conclusion:** A methodology for assessing the rescue benefit of deployed AEDs was proposed, and deployment strategies for maximizing the rescue benefit of AEDs along a long, narrow, corridor-like, finite space were derived. The strategies are simple and readily implementable. Our methodology can be easily generalized to search for optimal deployment of AEDs in planar areas or three-dimensional spaces.

## Introduction

Sudden cardiac arrest is a major public health issue worldwide (1, 2) and accounts for up to 20% of deaths in Western societies (3, 4). Out-of-hospital cardiac arrest (OHCA) describes an event of cardiac arrest before the patient is transferred to a hospital; that is, the cardiac arrest could occur in the field, in the community, at the patient’s home or workplace (5, 6). The first and second most common locations for an OHCA were at a residence and in public, respectively (7). OHCAs are witnessed by a layperson in 37% of cases and by an emergency medical services (EMS) provider in only 12% of cases (7). The survival rate after an OHCA is very low; at least 90% to 95% of these individuals do not survive despite resuscitation attempts (8). Evidence indicates that when the OHCA is caused by ventricular tachycardia or fibrillation, defibrillation is an effective treatment (9); however, its effectiveness diminishes with each passing minute (5, 10).

Public access to automated external defibrillators (AEDs) plays a key role in increasing the survival of patients with OHCA who have a shockable cardiac rhythm (11-13). Although AEDs are widespread, OHCAs defibrillated by bystanders before EMS arrival remains low, only approximately 2-4% (14). Therefore, to achieve the maximum benefit of public AEDs, careful evaluation of AED locations is required. While mathematical optimization has been used for assessing optimal locations for the AED deployment (15-19), most of these studies focus on numerical analysis/simulation of retrospective data. To our knowledge, few studies, if any, had taken an analytical approach to derive generalizable rules or strategies for the optimal AED deployment.

In this study, we aimed to explore such strategies for a finite one-dimensional space, which is a suitable model for long strip-like locations, such as trains, platforms in a train station, long tunnels, etc. By using simplified yet realistic assumptions, we derived simple rules for the optimal AED deployment that would maximize AED accessibility within a specified time frame.

## Methods

A location where an AED is placed is deemed “optimal” if the AED can be accessed within a specific time when OHCA occurs. Two essential factors need to be considered for assessing optimal AED deployment: time to acquire the AED and the probability of occurrence of OHCAs. The first factor leads to the notion of effective AED coverage.

### Effective coverage of an AED

- The sooner an AED is applied, the higher the survival chance of the OHCA (5, 10). According to timeliness of AED availability, the areas surrounding the AED are differentiated into three zones: hot, cold, and warm.
- Hot Zone: This is a region where an AED can be acquired to provide the best survival chance. According to the American Heart Association’s suggestion, the AED needs to be applied to patients with OHCA within one minute (7). An OHCA event is said to be within *the effective coverage* of an AED if it occurs within the hot zone of the AED.
- Cold Zone: This is a region where it takes longer time to obtain an AED than waiting for the EMS to arrive. An AED is ineffective in terms of the rescue benefit in its cold zone.
- Warm Zone: The is a region where an AED can be acquired before the EMS arrives but not within one minute. An AED provides some but not the best rescue in its warm zone.

For example, in our city, the EMS arrives at an OHCA event in eight minutes on average. Therefore, we set the cold zone to be where the time of AED arrival is more than eight minutes, and the warm zone is between one and eight minutes.

We assume that the AED moves with constant speed through the areas of interest. Thus, the *distance* between the AED and the OHCA event would decide the rescue benefit of the AED. The three zones surrounding the AED are then characterized by physical *distance* rather than *time*, as described above.

When more than one AED is under consideration, the combined hot zone of the collective AEDs is the *union* of the hot zones of each individual AED because it is evident that at least one AED can be acquired within 1 minute for the patient with OHCA in this region. Similarly, the combined cold zone of the collective AEDs is the *intersection* of all cold zones, as none of the AEDs can be acquired within 8 minutes of time in this region. Everywhere else is the combined warm zone, where none of the AEDs can be acquired within 1 minute, but at least one can be acquired between 1 and 8 minutes.

### Probability of OHCA occurrence

Intuitively, the larger the hot zone of an AED is, the more rescue benefit it provides. However, this is not entirely true. Evidently, an AED would provide no benefit if OHCA events never occur in its hot and warm zones. Furthermore, an AED would provide more benefit than another if more OHCA events occur in its hot zone. Thus, the “probability of OHCA occurrence per unit area” in hot/warm/cold zones of an AED needs to be considered when evaluating its rescue benefit. This quantity shall be used as a weight that indicates the relative importance of different areas surrounding an AED. The probability density of OHCA events in an area may vary with respect to, for example, population density, age distribution, history of heart disease, etc. (12).

### Evaluation of rescue benefit of AEDs

In this study, the rescue benefit of one or several AEDs is measured by the “weighted” sizes of the corresponding hot zone, cold zone, and warm zone. Let the area of interest be divided into standard unit areas, and let

- 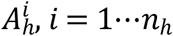, denotes the area units that constitute the hot zone;
- 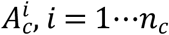 denotes the area units that constitute the cold zone;
- 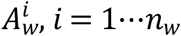 denotes the area units that constitute the warm zone;
- 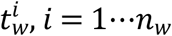 denotes the least amount of time (round up to the minute) for one AED to be acquired by an OHCA patient in the area unit 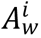;
- *dA* denotes the size of the unit area;
- p_*OHCA*_(·) denotes the probability density function of OHCA events over the area of interest.

Then, the weighted sizes of the hot zone, cold zone, and warm zone, denoted by *A*_*H*_, *A*_*C*_, *A*_*W*_, respectively, are computed as follows:

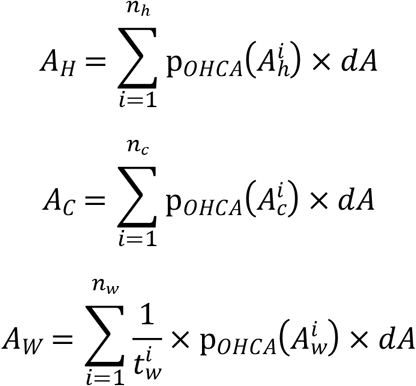

The rescue benefits of two sets of AEDs are assessed and compared by the following rules: one set of AEDs (S_AED1_) has a better rescue benefit than another set (S_AED2_)

1. if the weighted size of the hot zone *A*_*H*_ of S_AED1_ is *larger than* that of S_AED2_; or
2. if their respective *A*_*H*_ are the same and the weighed size of the cold zone *A*_*C*_ of S_AED1_ is *smaller than* that of S_AED2_; or
3. if their respective *A*_*H*_ and *A*_*C*_ are both the same and the weighed size of the warm zone *A*_*W*_ of S_AED1_ is *larger than* that of S_AED2_.

Should the two sets of AEDs have the same *A*_*H*_, *A*_*C*_, and *A*_*W*_, then their rescue benefits are regarded the same.

### Setup for the optimal AED deployment problem under consideration

In this study, we aimed to find a strategy for optimally deploying one or multiple AEDs along a long and narrow space, which can be mathematically modeled as a finite one-dimensional line segment. Such a model is suitable for real-life public spaces such as trains, platforms in train stations, tunnels, etc. The following assumptions are made:

**Assumption 1**. The geometric conditions of the space under consideration are uniform, and the AEDs can be transported along the space at a constant speed.

A**ssumption 2**. The probabilities for an OHCA event to occur at any two points in the space under consideration are identical.

The first assumption allows the hot, cold, and warm zones of an AED to be characterized by the distance to the AED, which, although simplifying, is reasonable when the space under consideration is of small to moderate size. The second assumption is reasonable when the space under consideration is public, and there is no reason to expect that a particular population with high OHCA risks would be present at specific locations. Note that by the second assumption, the “weighted sizes” of the hot and cold zones become the “actual sizes”, and the size of the warm zone becomes only inversely time-scaled. As an illustration, consider the spaces depicted in Fig. 1, where we assume it takes one minute to move through one unit area. Comparing the weighted sizes of the hot, cold, and warm zones, the two AEDs in Fig. 1A have the same rescue benefit, while the two sets of AEDs in Fig. 1B do not.

**Figure 1:**
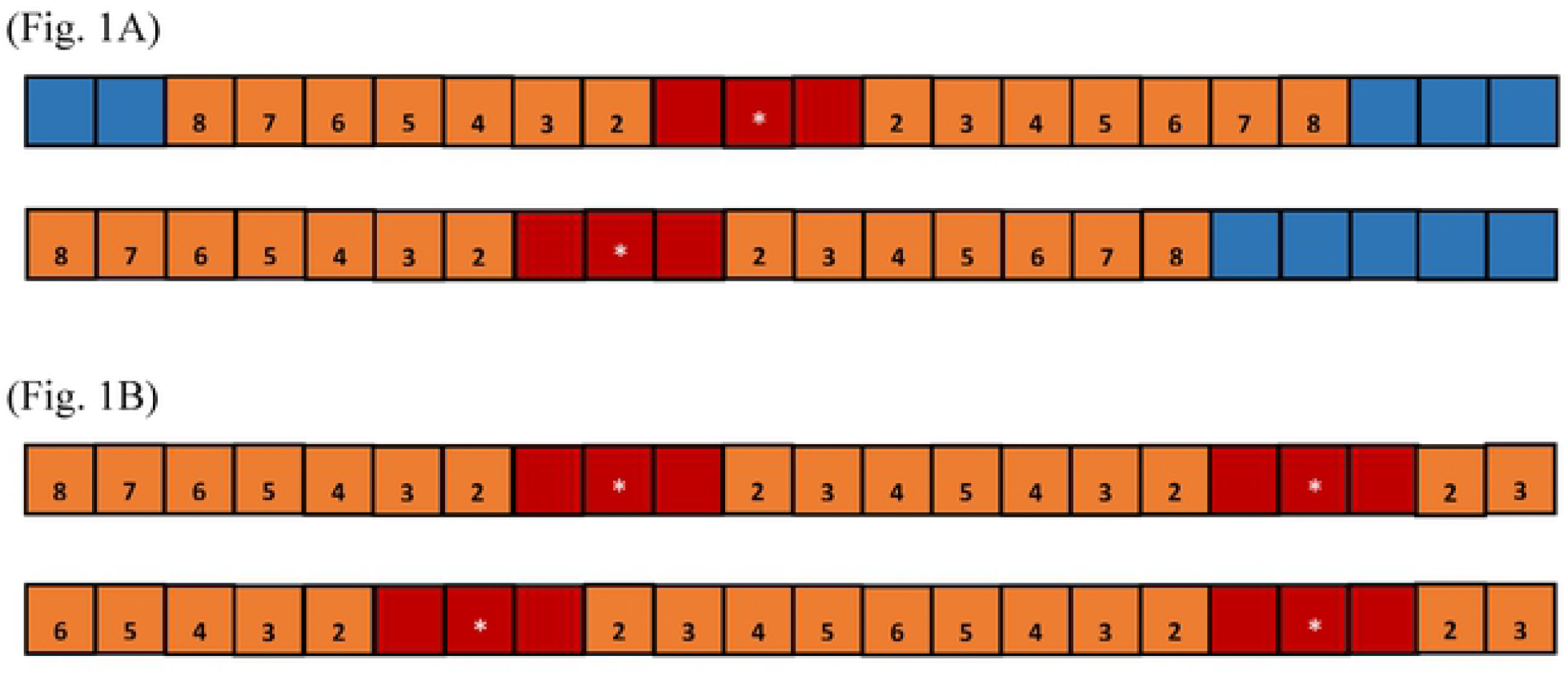
Illustration of weighted sizes of the hot, warm, and cold zones of one AED and two AEDs. (Fig. 1A. top) An AED is placed at the 11^th^ square from the left. The hot, warm, and cold zones are colored red, orange, and blue, respectively. For this example, the weighted sizes are *A*_*H*_ = 3, *A*_*c*_ = 5, and 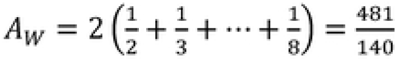. (Fig. 1A. bottom) If we place the AED at the 9^th^ square, the weighted sizes of hot. warm, and cold zones remain the same. The rescue benefit of this location is the same as that of the previous one. (Fig. 1B. top) An AED is placed at the 9^th^ square from the left, and another AED is placed at the 4^th^ square from the right. The hot and warm zones are colored red and orange, respectively. For this example. there is no cold zone, and the weighted sizes of the hot and warm zones arc *A*_*H*_ = 6 and 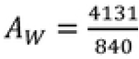. (Fig. 1B. bottom) If we move the left AED to the 7^th^ square, the weighted sizes of hot and warm zones become different, with *A*_*H*_ = 6 and 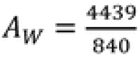. Therefore, moving the AEDs to the 7^th^ square results in a belter rescue benefit.

According to their accessibility, we consider three types of AEDs: fixed, summonable, and patrolling (subsequently denoted by F-type, S-type, and P-type, respectively).

- A fixed AED is an AED placed in a fixed location.
- A summonable AED is an AED placed in a location with a person on call to deliver the AED when receiving requests. We assume that the time delay between the occurrence of an OHCA and a summonable AED responding to the event is negligible.
- A patrolling AED is an AED carried by a person moving over a designated area periodically, who can deliver the AED when receiving requests.

Examples of summonable AEDs and patrolling AEDs are not uncommon. AEDs in the dining car of a train where someone can respond to a call to deliver the AEDs are summonable. AEDs carried by a person, such as a conductor or a service attendant walking through a train, become patrolling. The accessibility of these three types of AEDs drastically changes their corresponding *A*_*H*_, *A*_*C*_, and *A*_*W*_. Clearly, the *A*_*H*_ and *A*_*W*_ of a summonable AED are twice as large as those of a fixed AED. This is because a summonable AED is delivered on call, and therefore, the time for retrieving a summonable AED is half as much as that of a fixed AED at the same location. For a patrolling AED, the corresponding *A*_*H*_, *A*_*C*_, and *A*_*W*_ are evaluated as the average of those corresponding to a summonable AED placed in every area in the region it patrols. See Fig. 2 for an illustration of the hot, cold, and warm zones of these three types of AEDs.

**Figure 2:**
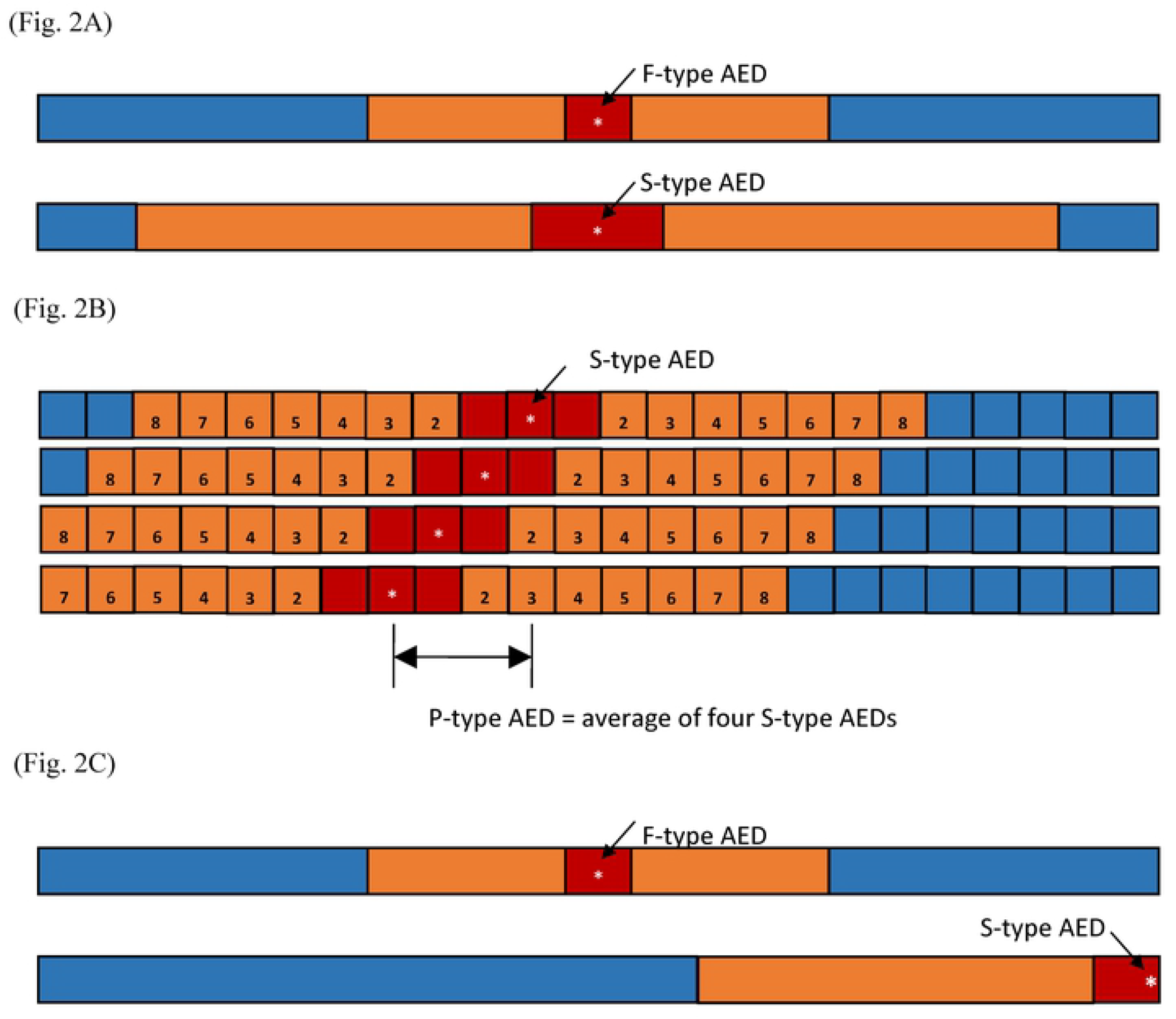
Illustration of the three types of AEDs: fixed, summonable, and patrolling. (Fig. 2A) The hot. warm, and cold zones of fixed (F-typc) AEDs and summonable (S-type) AEDs. The weighted sizes of the hot and warm zones *(A*_*H*_ and *A*_*W*_, respectively) of an S-type AED are twice as large as their counterparts of an F-type AED. As such, the cold zone of an S-type AED is smaller than that of an F-typc AED in the same confined space. (Fig. 2B) The weighted sizes of the hot, warm, and cold zones of a patrolling (P-type) AED. Assume that the P-type AED is patrolling between the 8^th^ and 11^th^ (from the left) squares. The weighted sizes *A*_*H*_, *A*_*c*_. and *A*_*W*_ of this P-type AED arc the averages of those corresponding to the four S-type AEDs placed at the 8^th^ to 11^th^ squares, respectively. (Fig. 2C) The *A*_*H*_ and *A*_*W*_ of an S-type AED are twice as large as those of an F-type AED. Thus, even when an S-type AED is placed at the least optimal position where its *A*_*H*_ and *A*_*W*_ are halved, the rescue benefit it provides is still the same as that of an F-type AED placed at the optimal position.

Rules for optimally deploying the AEDs are given in the next section. The rules were derived mathematically and numerically verified by computer programs. The numerical verification is performed based on dividing the one-dimensional space of interest into a collection of finite unit areas and searching through all possible deployments to determine the optimal deployments, which are determined by comparing the rescue benefits as discussed previously. To accommodate the numerical computation and all practical purposes regarding the applications we have in mind, we consider up to five AEDs in our numerical verifications, with only one patrolling AED. We assume that the patrolling AED moves through the whole space.

## Results

The rules for optimally deploying the AEDs are presented here. The rules are given for one, two, and multiple (up to five) AEDs of the three types discussed in the previous section.

### Optimal deployment strategy for one AED

Given a finite one-dimensional space, a fixed AED would provide the best rescue benefit if its hot and warm zones are fully contained in the space, as this would result in its cold zone being minimized. Furthermore, two fixed AEDs would have the same rescue benefit if their respective hot zones and warm zones are fully contained in the space. Finally, as illustrated in Fig. 2C, placing a fixed AED toward either side of the finite space may result in parts of its hot and warm zones falling outside the space and thus enlarging the cold zone. Following these arguments, the optimal AED location is in *the center* of the space. The worst AED locations are the two end points. The same arguments are also applied to a summonable AED.

Moreover, as the *A*_*H*_ and *A*_*W*_ of a summonable AED are twice as large as those of a fixed AED and placing an AED at the end of the space would reduce its *A*_*H*_ and *A*_*W*_ by half, one concludes that placing a summonable AED at the worst location would still have the same rescue benefit as a fixed AED at its optimal location. Therefore, the benefit of a summonable AED is always larger than or equal to that of a fixed AED.

For AED patrolling throughout the whole space, its instantaneous benefit decreases as it moves toward the end point. Since the effective *A*_*H*_ and *A*_*W*_ of a patrolling AED is calculated in the averaged sense, as illustrated in Fig. 2B, the benefit of a patrolling AED is between the best and the worst rescue benefit that a summonable AED can provide.

### Optimal deployment strategy for two AEDs

When there are two AEDs, the benefit is maximized if the hot and warm zones of one AED do not intersect with their counterparts of the other AED; as in this case, the combined hot zone is maximized and the combined cold zone is minimized. Because placing an AED toward the “center” is potentially better, an optimal deployment strategy for two non-patrolling AEDs can be derived as follows: divide the whole space into two segments according to the ratio of the weighted sizes of the individual hot zones and place the AEDs at the respective centers of these segments. In the case of two fixed AEDs or two summonable AEDs, the space is to be divided into two equal segments. In the case of one fixed AED and one summonable AED, the space is divided into two segments, of which the ratio of the lengths is 1-to-2. See Fig. 3A for an illustration.

**Figure 3:**
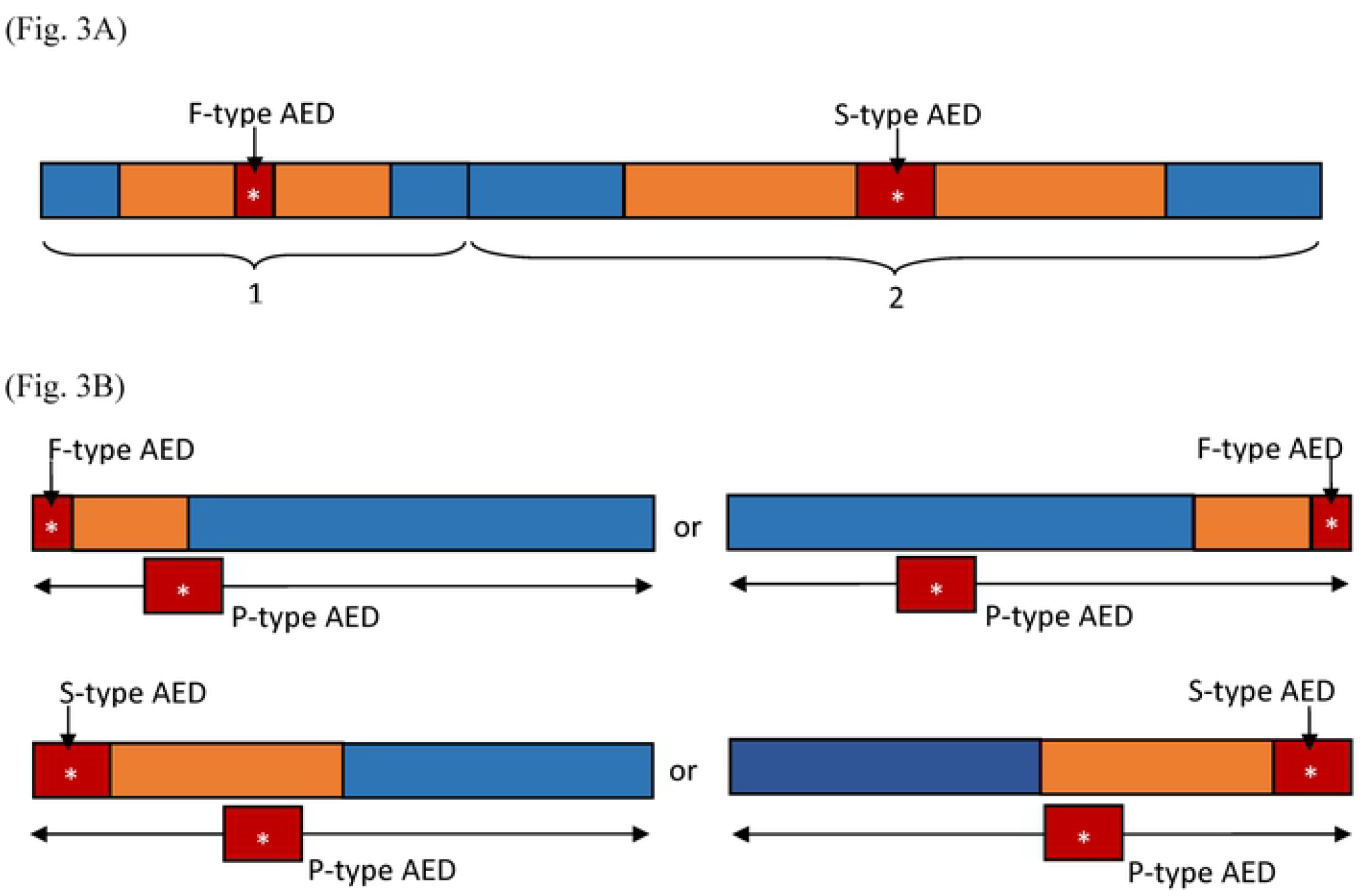
Illustration of the optimal deployment for two AEDs. (Fig. 3A) One F-type and one S-type AED. In this ease, one should divide the confined space into two parts with a l-to-2 length ratio. The F-type AED is placed at the center of the shorter part, and the S-type AED is placed at the center of the longer part. (Fig. 3B) One P-type AED and one F-type or S-type AED. In this case, the F-type or S-typc AED is placed toward one of the end locations with its hot zone entirely contained in the confined space. The P-typc AED patrols the entire space.

When one of the AEDs is patrolling throughout the space, the optimal location to place the non-patrolling AED is somewhat different from the “centering” strategy. The optimal location for placing the non-patrolling AED is near one of the end points of the space. Specifically, the fixed or summonable AED should be placed as close to the end point as possible, such that its hot zone is entirely within the space with its edge coinciding with the edge of the space. See Fig. 3B for an illustration.

### Optimal deployment strategy for multiple (three or more) AEDs

The idea developed in the previous sections can be generalized for optimal deployment of three or more AEDs. The general rules are given as follows.

- (Rule NP) In the case of no patrolling AED, the total space is divided into the sum of *n*_*F*_ shorter equal-length segments and *n*_*S*_ longer equal-length segments, with the length ratio of the longer to shorter segments being 2-to-1, where *n*_*F*_ and *n*_*S*_ are the numbers of fixed AEDs and summonable AEDs, respectively. Then, the fixed AEDs are placed at the center of the shorter segments, while the summonable AEDs are at the center of the longer segments. See Fig. 4 for an illustration.

**Figure 4:**
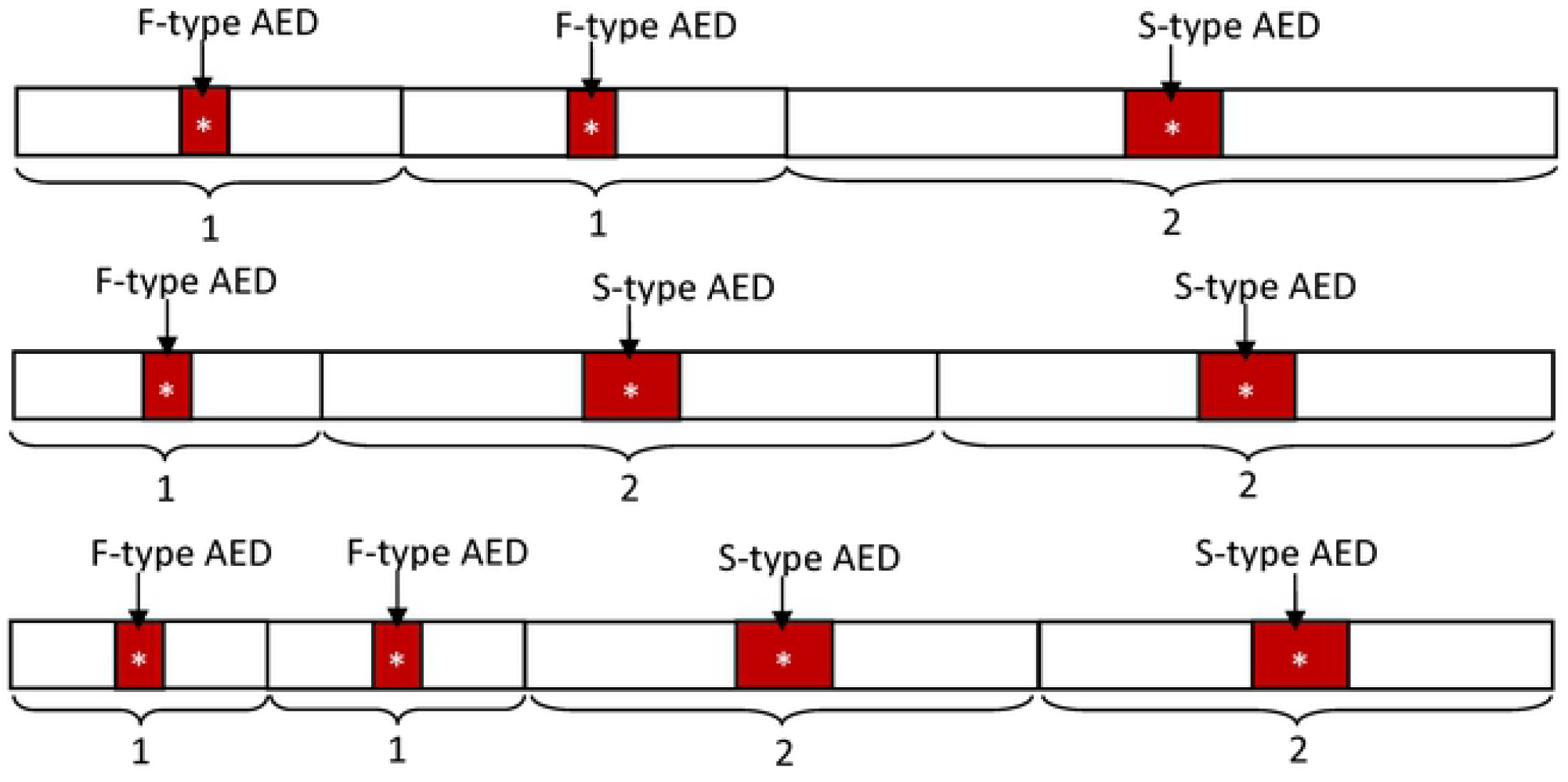
Illustration of optimal deployment for multiple AEDs. *n*_*F*_ F-type AEDs and *n*_*s*_ S-type AEDs, where (*n*_*F*_, *n*_*s*_) = (2, 1), (1, 2), and (2, 2). Here we only indicate the hot zones of each AEDs. In the case of (*n*_*F*_, *n*_*s*_) = (2, 1), the confined space is divided into three parts with a 1 -to-1 -to-2 length ratio. The F-type AEDs are placed at the centers of the shorter parts, and S-type AEDs are the longer parts. Similarly, in the case of (*n*_*F*_, *n*_*s*_) = (1, 2). the confined space is divided into three parts with a l-to-2-to-2 length ratio. In the case of (*n*_*F*_, *n*_*s*_) = (2, 2),. four parts are needed with a length ratio of l-to-l-to-2-to-2.
- (Rule WP) In the case of AEDs patrolling throughout the space, first choose two AEDs with the smallest individual hot zone and place them near the two ends of the space, such that the hot zones are entirely within the space and the edges of their hot zones coincide with the edges of the space. Then, follow Rule NP to place the remaining AEDs in the space excluding the two hot zones. Fig. 5 illustrates the idea.

**Figure 5:**
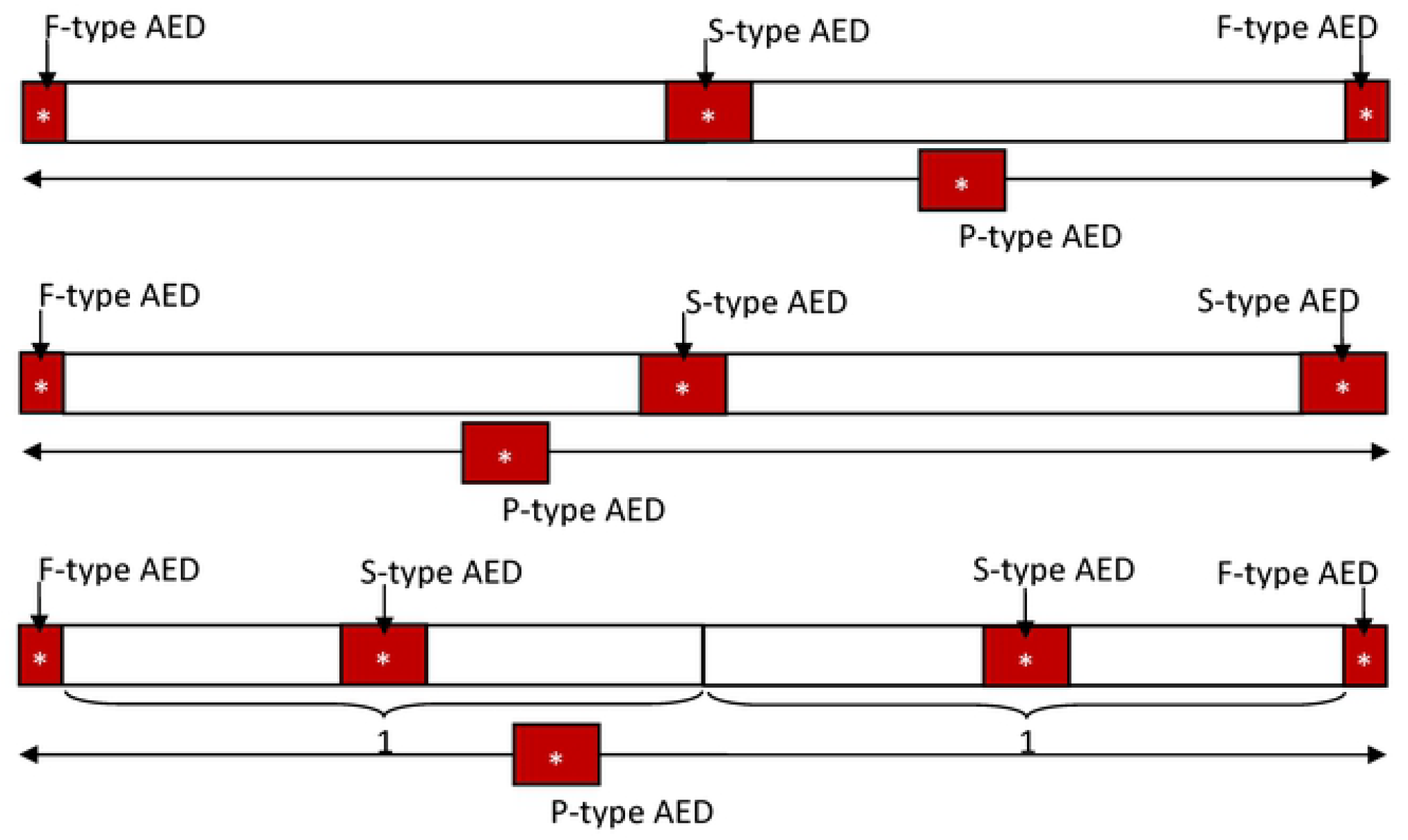
Illustration of optimal deployment for multiple AEDs. *n*_*F*_ F-type AEDs and *n*_*s*_ S-type AEDs with one patrolling (P-type) AED. where (*n*_F_, *n*_s_) = (2, 1), (1, 2), and (2,2). Again, here, we only indicate the hot zones of each AED. With a P-type AED, we need to place two AEDs toward the two end locations with their hot zones entirely contained in the confined space. In the case of (*n*_*F*_, *n*_*s*_) = (2, 1). the two F-type AEDs are placed toward the end locations, and the S-type AED is placed at the center of the remaining space. In the case of (*n*_*F*_, *n*_*s*_) = (1, 2), we place one F-type AED and one S-type AED toward the end locations and one S-type AED at the center of the remaining space. In the case of (*n*_*F*_, *n*_*s*_) = (2,2), again, we place two F-type AEDs toward the end locations. The remaining space is then divided into two parts with a 1 -to-1 length ratio. The two S-type AEDs arc then placed at the center of these two parts.

The optimality of these rules is verified numerically by computer programs for instances (*n*_*F*_, *n*_*S*_) = 2, 1), (1, 2), and (2, 2), with or without a patrolling AED.

The rules are summarized in Table 1.

**Table 1.** Summary of the optimal deployment of one, two, and three or more AEDs in a one-dimensional space.

| Number of AEDs | Type of AED | Deployment Rule for Maximizing Rescue Benefit |
| --- | --- | --- |
| one | F-type | Place the AED at the center of the finite space. |
|  | S-type | Place the AED at the center of the finite space. |
|  | P-type | The AED constantly patrols the whole space. |
| two | F-type + F-type or<br>S-type + S-type | Divide the finite space into two equal segments, and place one AED at the center of each segment, respectively. |
|  | F-type + S-type | Divide the finite space into two segments by length ratio 1-to-2 and place the F-type AED at the center of the shorter segment, and the S-type AED at the center of the longer segment. |
|  | F-type + P-type or<br>S-type + P-type | Place the F-type or the S-type AED at one end of the space so that the edge of its hot zone coincides with the edge of the space. The P-type AED constantly patrols the whole space. |
|  | P-type + P-type | The AEDs constantly patrol the whole space |
| three or more | F-type and/or S-type, no P-type | Divide the finite space into $n_F$ shorter segments plus $n_S$ longer segments, where $n_F$ is the number of F-type AEDs and $n_S$ is the number of S-type AEDs. The length ratio between the shorter and longer segments is 1-to-2. Then, place one F-type AED at the center of the shorter segment and one S-type AED at the center of longer segment. |
|  | at least one P-type, together with either F-type and/or S-type | The P-type AED(s) constantly patrols the whole space. At each end of the space, place one F-type or S-type AED so that the edge of its hot zone coincides with the edge of the space. Finally, deploy the remaining F-type and S-type AEDs to the space excluding the two hot zones according to the rule stated above. |

## Discussion

Given several AEDs, to maximize the overall rescue benefit of these AEDs, evaluated by the weighted size of their collective hot zones, one should deploy the AEDs such that the hot zone of one AED avoids overlapping with those of the other AEDs as much as possible. This is the central principle behind all the rules presented in the previous section. Note that the rules we presented are not the only way to maximize the rescue benefit, but they are simple and easy to implement.

The results we derived are typically applicable to public spaces such as trains and tunnels. They are also applicable to tall buildings when the vertical coverage of AEDs across stories is concerned. In such scenarios, a tall building can be effectively viewed as a finite one-dimensional space.

Our results were derived under the assumption that the geometric conditions of the space under consideration are uniform, that the patrolling AEDs periodically travel the whole space with constant speed, and they are transported along the space with a constant speed. We also assumed that the probabilities for an OHCA event to occur at any point in the space are identical. These simplifying assumptions, which are realistic to some applications as explained, allowed us to mathematically derive simple rules for optimal deployment. In cases where these assumptions do not hold, the methodology we proposed is still applicable as long as relevant parameters are known for evaluating the weighted sizes of hot, cold, and warm zones of the AEDs. The corresponding optimal deployment problem can still be solved numerically by well-developed algorithms (20). Moreover, the methodology we proposed can be straightforwardly generalized to accommodate two-dimensional problems, which are relevant to, for example, deployment of AEDs across a metropolitan region, and three-dimensional problems, which are relevant to, for example, deployment of AEDs in a commercial district with many skyscrapers where the vertical coverage of AEDs is essential.

## Conclusions

A methodology for assessing the rescue benefit of deployed AEDs was proposed, and deployment strategies for maximizing the rescue benefit of the AEDs along a long and narrow space were derived. The strategies are simple and readily implementable. Our methodology can easily be generalized to search for optimal deployment of AEDs in planar areas or three-dimensional spaces.

## Data Availability

All relevant data are within the manuscript and its Supporting Information files.

## Acknowledgements

This study was funded by the Taiwan Ministry of Science and Technology (Grant numbers: MOST 105-2314-B-006-069, MOST 106-2314-B-006-003, MOST 107-2314-B-006-003, and MOST 109-2221-E-110-031).

